# Bayesian shared-component spatiotemporal modeling of sexually transmitted infection co-occurrence: identifying geographic vulnerability across 204 countries, 1990–2023

**DOI:** 10.64898/2026.07.19.26358422

**Authors:** Qiancheng Ma, Tianzhen Zhang, Wen Yuan Zou, Dongzhi Lin

**Affiliations:** Department of Community Health, Faculty of Medicine and Health Sciences, Universiti Putra Malaysia, Serdang, Malaysia; School of Nursing and Midwifery, Trinity College Dublin, Dublin, Ireland; Department of Nursing, Faculty of Medicine and Health Sciences, Universiti Putra Malaysia, Serdang, Malaysia; School of Information Engineering, Fujian Polytechnic of Water Conservancy and Electric Power

**Keywords:** STI co-occurrence, Bayesian shared-component model, spatial vulnerability, global epidemiology, integrated STI prevention, disease mapping

## Abstract

**Objectives:** Although HIV incidence has declined in some settings, the overall global burden of sexually transmitted infections remains a major public health concern. In the context of the World Health Organization’s call for people-centred STI prevention and care, identifying the shared geographic pattern of multiple STIs using data-driven analysis may help detect vulnerable areas and inform integrated prevention strategies.

**Methods:** We analysed country-level incidence counts from the Global Burden of Disease 2023 study for 204 countries and territories over 1990–2023. A Bayesian shared-component spatiotemporal model was fitted, decomposing each disease’s log-rate into a shared spatial component (scaled intrinsic conditional autoregressive prior), disease-specific spatial deviations, disease-specific first-order random walk temporal effects, and five socioeconomic covariates, with a negative binomial likelihood to accommodate overdispersion. The shared spatial score—the posterior mean of the shared spatial component—was used as a continuous index of STI co-occurrence burden. Posterior exceedance probabilities quantified directional stability. External validity was assessed via Spearman correlation with the Socio-demographic Index and generalised estimating equation regression of HIV/AIDS mortality on the shared score.

**Results:** The shared spatial score exhibited marked geographic heterogeneity. The five highest-scoring countries were Eswatini (2.25), Lesotho (2.13), Malawi (1.90), Mozambique (1.89), and South Africa (1.85), all in southern Africa. Fifty-seven countries had high directional stability (posterior exceedance probability >0.95), concentrated in sub-Saharan Africa and the Caribbean. The score correlated negatively with SDI (Spearman ρ = −0.619, p = 6.4 × 10□^23^) and positively with HIV/AIDS mortality (incidence rate ratio = 14.64 per standard deviation, 95% CI: 11.90–18.01). Prior sensitivity analysis confirmed near-perfect ranking stability (ρ ≥ 0.9999).

**Conclusions:** STI co-occurrence is geographically concentrated, with the highest shared burden in sub-Saharan Africa and persistently elevated shared spatial signals also observed in parts of mainland Southeast Asia and the Caribbean. The shared spatial score provides a data-driven tool for prioritising integrated STI screening and prevention resources across countries.

## 1. Introduction

Sexually transmitted infections (STIs) remain a major global public health concern because of their substantial and persistent disease burden worldwide. According to the World Health Organization, more than 1 million sexually transmitted infections are acquired every day, indicating that STI transmission continues at a very large scale globally[1]. In response to this ongoing burden, the WHO has set targets for 2030 to reduce the impact of STIs as a public health problem[2]. In addition, findings from the Global Burden of Disease (GBD) study have consistently shown that the burden of HIV and other sexually transmitted infections remains considerable at the global level, further highlighting the sustained public health importance of STIs(see Fig 1 for more detils)[3].

**Fig 1.**
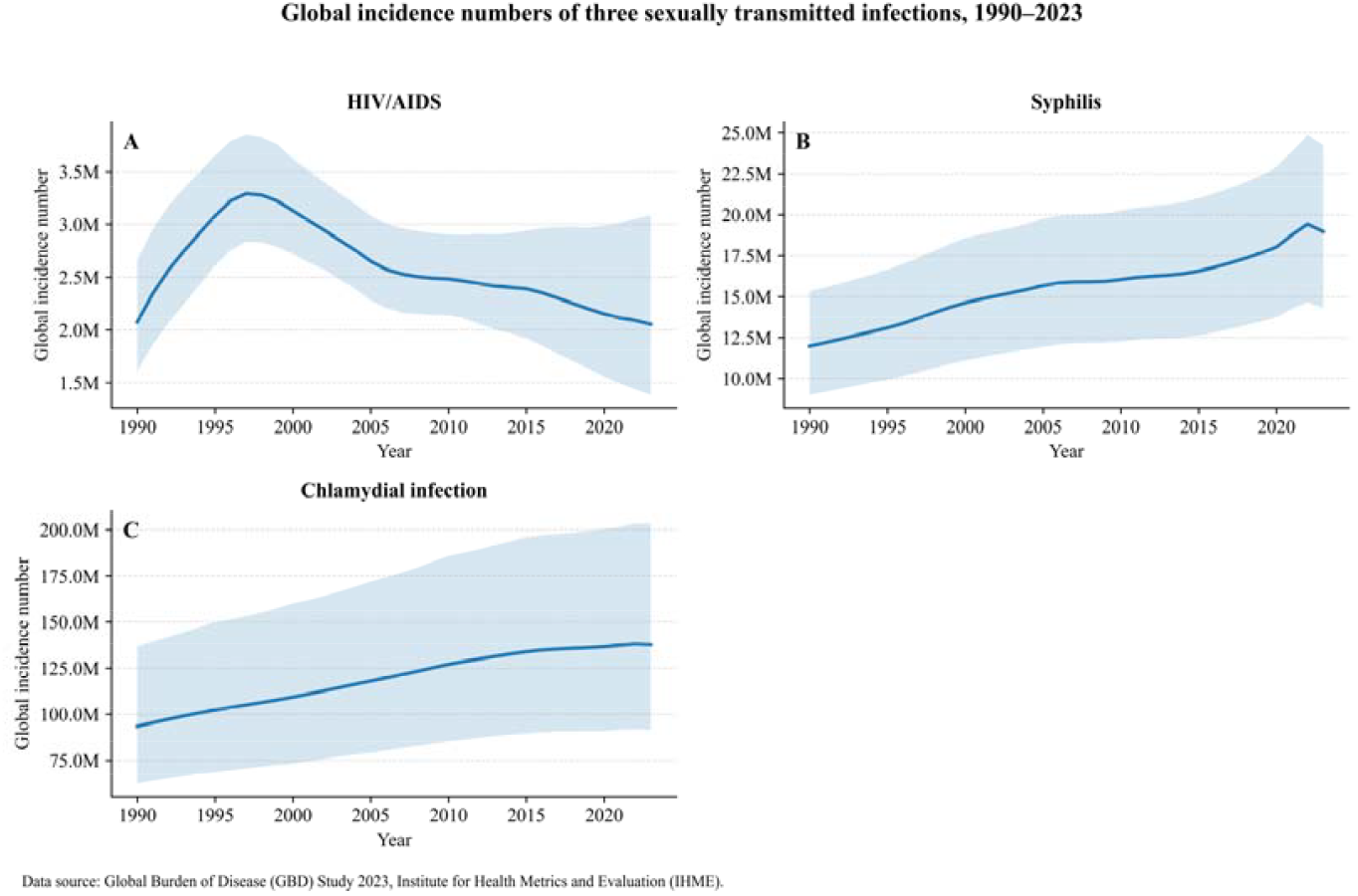
Global incidence number of three sexually transmitted infection,1990-2023 (Note: The figures were created using Python.)

Investigating STI co-occurrence is important because it may reveal underlying vulnerabilities in local sexual health prevention and control. In areas where sexual health awareness is limited, people may have insufficient knowledge of STI prevention, early detection, and treatment, which can increase the risk of ongoing transmission of multiple infections. Moreover, some STIs may facilitate the transmission of others through biological synergy. For instance, infections such as syphilis or genital warts may disrupt mucosal or epithelial integrity and increase exposure to infectious body fluids during sexual activity, thereby creating conditions that favor further pathogen transmission[4, 5]. From a public health perspective, the geographic clustering of multiple STIs may therefore reflect not only shared transmission risks, but also weaknesses in local prevention networks, including insufficient education, screening, treatment, and sexual health services.

Existing spatial research on sexually transmitted infections still has important limitations. Most previous studies have focused on the spatial clustering of a single STI, with the main emphasis placed on describing the geographic distribution, hotspot areas, or regional disparities of one disease at a time[6]. Even when multiple STIs have been examined, they have often been presented in parallel using maps to describe the spatial patterns of each infection separately, rather than using statistical approaches to test whether these diseases share a stable underlying spatial pattern[7]. As a result, although the existing literature may suggest that several STIs tend to concentrate in the same areas, it remains unclear whether such overlap reflects a true cross-disease spatial co-occurrence pattern or merely a descriptive geographic coincidence. Evidence is even more limited from a global perspective, with a particular lack of model-based analyses examining the spatial co-occurrence of multiple STIs across countries. At the same time, the World Health Organization has increasingly emphasized integrated, people-centred STI services as part of the response to the global STI burden[8, 9]. However, there is still limited global quantitative evidence to identify which areas should be prioritized for integrated STI prevention and control efforts.

This study aimed to identify spatial patterns that may be shared across multiple sexually transmitted infections at the global level. Specifically, we used a Bayesian shared-component spatiotemporal model to jointly analyse incidence data for HIV/AIDS, syphilis, and chlamydial infection, in order to identify consistent cross-disease spatial co-occurrence hotspots rather than simply describing the geographic distribution of each disease separately. On this basis, we further evaluated the epidemiological plausibility of the identified hotspot pattern through external validation (HIV mortality), to assess whether it may reflect a form of shared geographic vulnerability with public health relevance. Ultimately, we aimed to provide a more quantitative evidence base for the geographic prioritisation of integrated STI prevention and control resources.

## 2. Methods

### 2.1. Study design and data sources

This was a global ecological spatiotemporal analysis using GBD 2023 data to examine STI spatial co-occurrence patterns across 204 countries from 1990 to 2023. The three STIs included were HIV/AIDS, syphilis, and chlamydial infection, and the outcome for all three diseases was incidence counts (all ages, both sexes). During the study period, the cumulative incidence counts were 88,314,816 for HIV/AIDS, 526,687,320 for syphilis, and 4,058,030,234 for chlamydial infection. Each disease contributed 6,936 country-year observations, corresponding to complete observations for 204 countries over 34 years. Covariates included the Socio-demographic Index (SDI) from GBD, as well as population density, health expenditure per capita, the Universal Health Coverage (UHC) service coverage index, and physician density from the World Bank. Spatial boundary data were obtained from the Natural Earth 10m shapefile and were used to construct the country-level spatial adjacency structure for subsequent spatiotemporal modelling.

### 2.2. Statistical model

We fitted a Bayesian shared-component spatiotemporal model to jointly analyse incidence counts of HIV/AIDS, syphilis, and chlamydial infection at the country-year level. Let *y*_*ikt*_ denote the observed incidence count for disease *k*in country *i* and year *t*. We assumed a negative binomial likelihood:

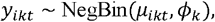

where *µ*_*ikt*_is the mean incidence count and *ϕ*_*k*_is the disease-specific overdispersion parameter. The logarithm of the expected count was modelled as:

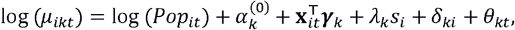

where *log* (*Pop*_*it*_)is the population offset; 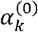 is the disease-specific intercept; **x**_*it*_ is the covariate vector with disease-specific coefficients ***γ***_*k*_; *s*_*i*_ is the shared spatial component; *λ*_*k*_is the disease-specific non-negative loading linking disease *k*to the shared spatial pattern; *δ*_*ki*_ is the disease-specific spatial deviation; and *θ*_*kt*_is the disease-specific temporal effect. Covariates included SDI, log-transformed population density, log-transformed health expenditure per capita, UHC service coverage index, and physician density per 1,000 population; all were z-score standardised prior to model fitting. Full specification of prior distributions, spatial structure (scaled ICAR with K-nearest neighbour fallback for isolated territories), temporal structure (non-centred RW(1) with sum-to-zero constraint), and MCMC sampling configuration are provided in Supplementary Text S1.

### 2.3. Model diagnostics and validation

Model performance was evaluated through five complementary diagnostic and validation procedures. First, convergence of the MCMC sampler was assessed via 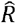, effective sample size (ESS), and the number of divergent transitions. Second, model fit was evaluated using LOO-CV and WAIC with Pareto *k* diagnostics, supplemented by posterior predictive checks. Third, prior sensitivity was assessed by re-fitting the model under two alternative prior configurations. Fourth, collinearity among covariates was examined using variance inflation factors (VIF) and Spearman correlation matrices. Fifth, the epidemiological plausibility of the identified hotspots was assessed through two external validation analyses: (i) convergent validity with SDI via Spearman rank correlation, and (ii) convergent validity with HIV/AIDS mortality via GEE negative binomial regression. Full details of each diagnostic procedure and associated results are provided in online supplementary Text S2.

### 2.4. Shared spatial score

The primary output of the model is the country-level shared spatial score, defined as the posterior mean of the shared spatial component:

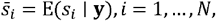

estimated as the Monte Carlo average over *S* = 32,000 posterior samples. This continuous index summarises the country-specific latent spatial pattern shared across HIV/AIDS, syphilis, and chlamydial infection after accounting for covariates, disease-specific spatial effects, and disease-specific temporal effects. Because the shared spatial component is centered, positive values indicate countries with higher-than-reference shared spatial tendency, whereas negative values indicate lower-than-reference shared spatial tendency; the magnitude reflects the strength of deviation on the log-rate scale.

### 2.5. Posterior exceedance probability

To quantify the uncertainty of the shared spatial score, we calculated the posterior exceedance probability for each country as

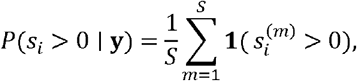

where 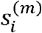 denotes the *m*-th posterior sample of the shared spatial component. This probability reflects the posterior support that a country’s shared spatial tendency is above the centered reference level. Higher values indicate stronger evidence that the country belongs to the shared hotspot pattern, whereas lower values indicate evidence toward a shared coldspot pattern.

### 2.6. External validation

#### 2.6.1. External validation 1: convergent validity with SDI

We evaluated whether the Shared spatial score was associated with national socioeconomic development by computing the Spearman rank correlation between the country-level Shared spatial score and SDI. Let *s*_*i*_ denote the Shared spatial score for country *i*, and let *SDI*_*i*_ denote the corresponding SDI value. The correlation of interest was

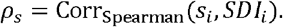

#### 2.6.2. External validation 2: convergent validity with HIV/AIDS mortality

We further assessed whether the Shared spatial score was associated with HIV/AIDS mortality using a generalized estimating equation with a negative binomial family and log link. Let *Y*_*it*_ denote the HIV/AIDS death count for country *i* in year *t*, and let *Pop*_*it*_ denote the corresponding population. The main model was specified as

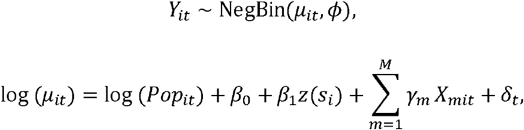

where *Y*_*it*_ denotes the HIV/AIDS death count for country *i* in year *t*; *µ*_*it*_is the expected mean count; *ϕ*is the dispersion parameter; *Pop*_*it*_ is the population used as the offset; *z*(*s*_*i*_) is the standardized shared spatial score; *X*_*mit*_ denotes the *m*-th covariate; *γ*_*m*_ is the corresponding regression coefficient for the *m*-th covariate; and *δ*_*t*_ represents year fixed effects.

As a sensitivity analysis, we also categorized countries into high and low shared-hotspot groups based on the upper and lower quartiles of the Shared spatial score distribution. The corresponding GEE model replaced the continuous Shared spatial score with a binary indicator:

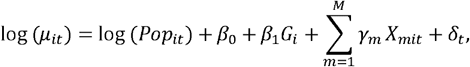

where *G*_*i*_ = 1 indicates membership in the upper-quartile shared-hotspot group and *G*_*i*_ = 0 indicates membership in the lower-quartile group.

## 3. Results

### 3.1. Model diagnostics

Detailed diagnostic procedures and full results are provided in **Supplementary Text S2**.

MCMC convergence was assessed via the potential scale reduction factor (R□), effective sample size (ESS), and the number of divergent transitions. All criteria were satisfied (see Table 1). These results confirm that the sampler thoroughly explored the posterior distribution.

**Table 1.**
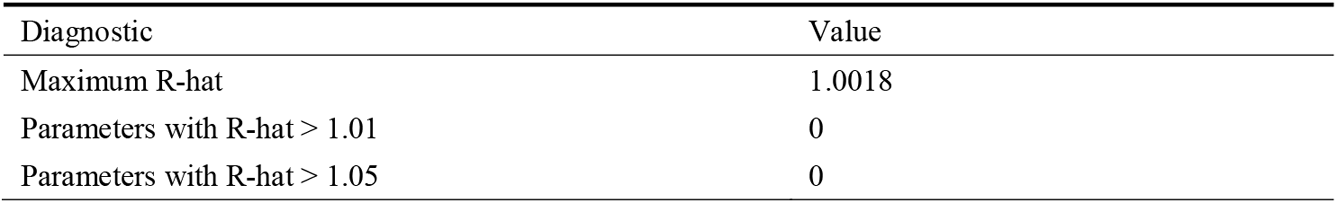
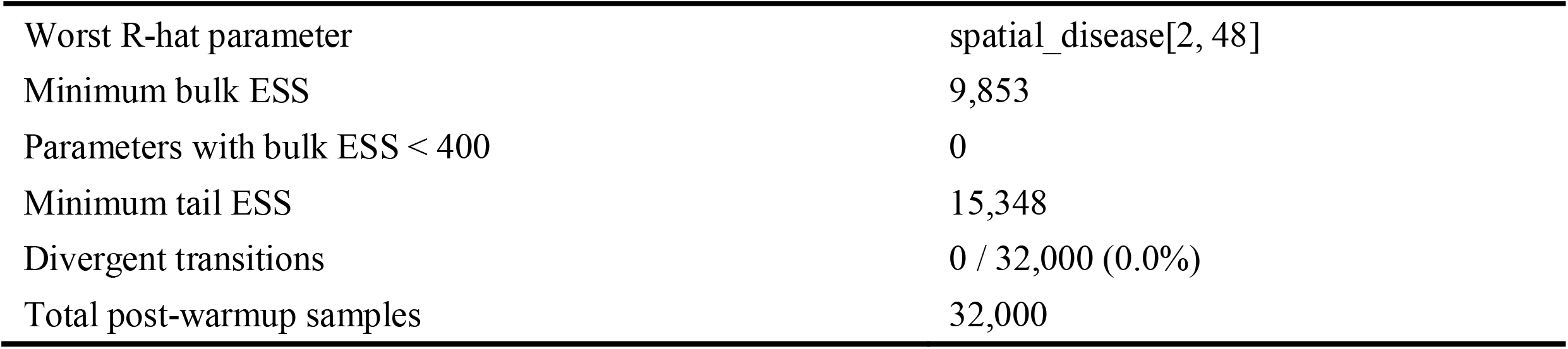
MCMC convergence summary.

Posterior predictive checks supported adequate model fit. The observed-versus-predicted scatter plot showed strong agreement along the diagonal with no systematic deviation (see Fig 2), and approximately 92.54% of observed values fell within the 90% posterior predictive intervals (see Fig 3).

**Fig 2.**
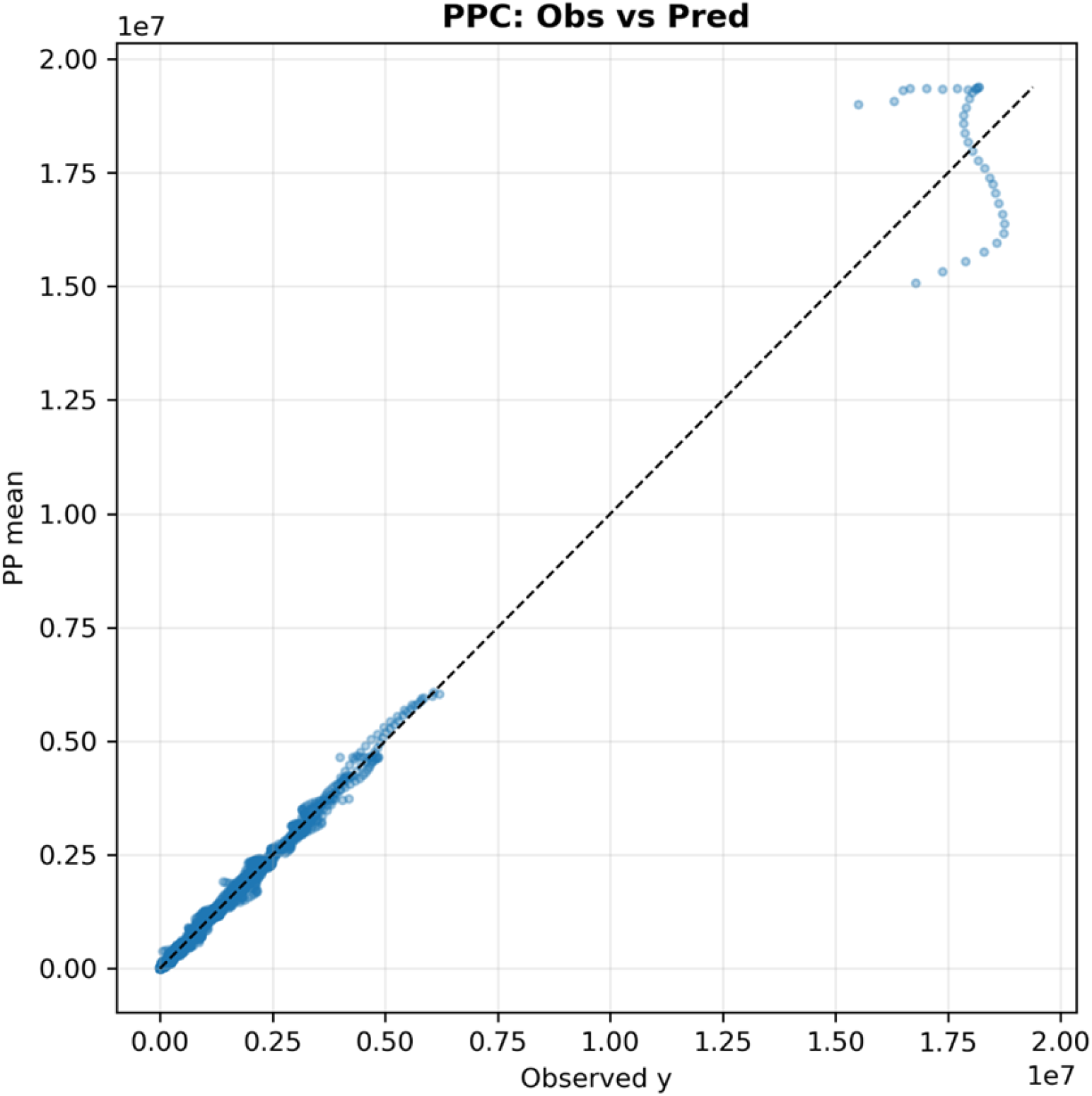
Observation-level posterior predictive check: observed values versus posterior predictive means (Note: The figures were created using Python.)

**Fig 3.**
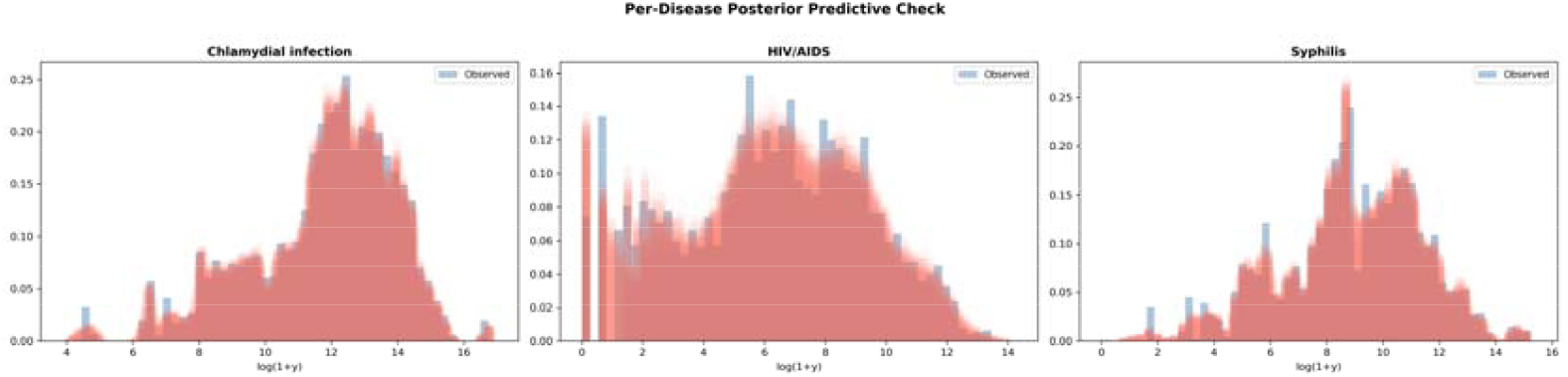
Per-disease posterior predictive check (Note: Blue histograms show the observed distribution of log (1 + y); red shaded areas show 50 replicated datasets drawn from the posterior predictive distribution. The figures were created using Python.)

Predictive fit diagnostics were consistent across criteria. The close agreement between LOO-CV and WAIC estimates indicates that the predictive evaluation was numerically stable and not sensitive to the choice of information criterion (see Table 2).

**Table 2.**
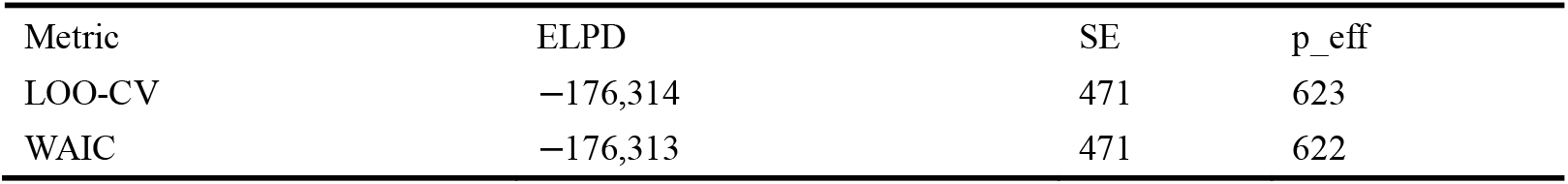
Model comparison via LOO-CV and WAIC.

All 20,808 observations had Pareto k ≤ 0.70, indicating that no single observation exerted undue influence on the model fit(see Table 3).

**Table 3.**
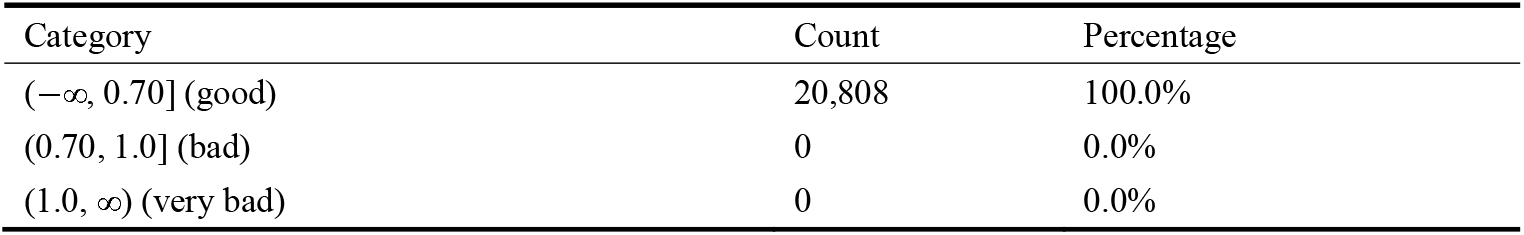
Pareto k diagnostics.

| Category | Count | Percentage |
| --- | --- | --- |
| $(-\infty, 0.70]$ (good) | 20,808 | 100.0% |
| $(0.70, 1.0]$ (bad) | 0 | 0.0% |
| $(1.0, \infty)$ (very bad) | 0 | 0.0% |

Prior sensitivity analysis confirmed that the shared spatial score rankings were robust to alternative prior specifications (see Table 4). The top-10 hotspot countries were identical across all configurations.

**Table 4.**
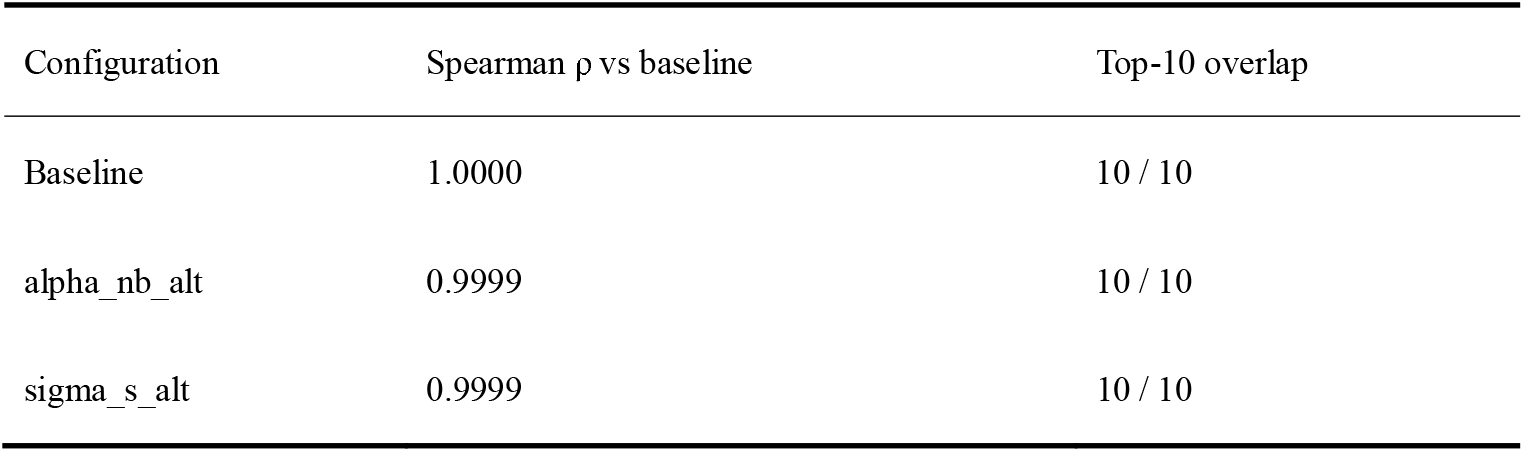
Prior sensitivity analysis results.

### 3.2. Global distribution of the shared spatial score

Global distribution of the shared spatial score. The shared spatial score showed substantial cross-country heterogeneity. The five highest-scoring countries were Eswatini (2.249), Lesotho (2.126), Malawi (1.903), Mozambique (1.891), and South Africa (1.846). More broadly, the top 50 countries were concentrated predominantly in sub-Saharan Africa, especially southern and eastern Africa, with additional representation from the Caribbean and only a small number of countries in Asia. In contrast, the five lowest-scoring countries were Tokelau (-6.118), Niue (-6.068), Cook Islands (-3.867), North Macedonia (-1.930), and Albania (-1.868). Low-score countries were mainly concentrated in Pacific island settings and southeastern Europe / the Balkans, with additional low-score countries observed in parts of the Middle East (see **Fig 4** and Supplementary Table S1).

**Fig 4.**
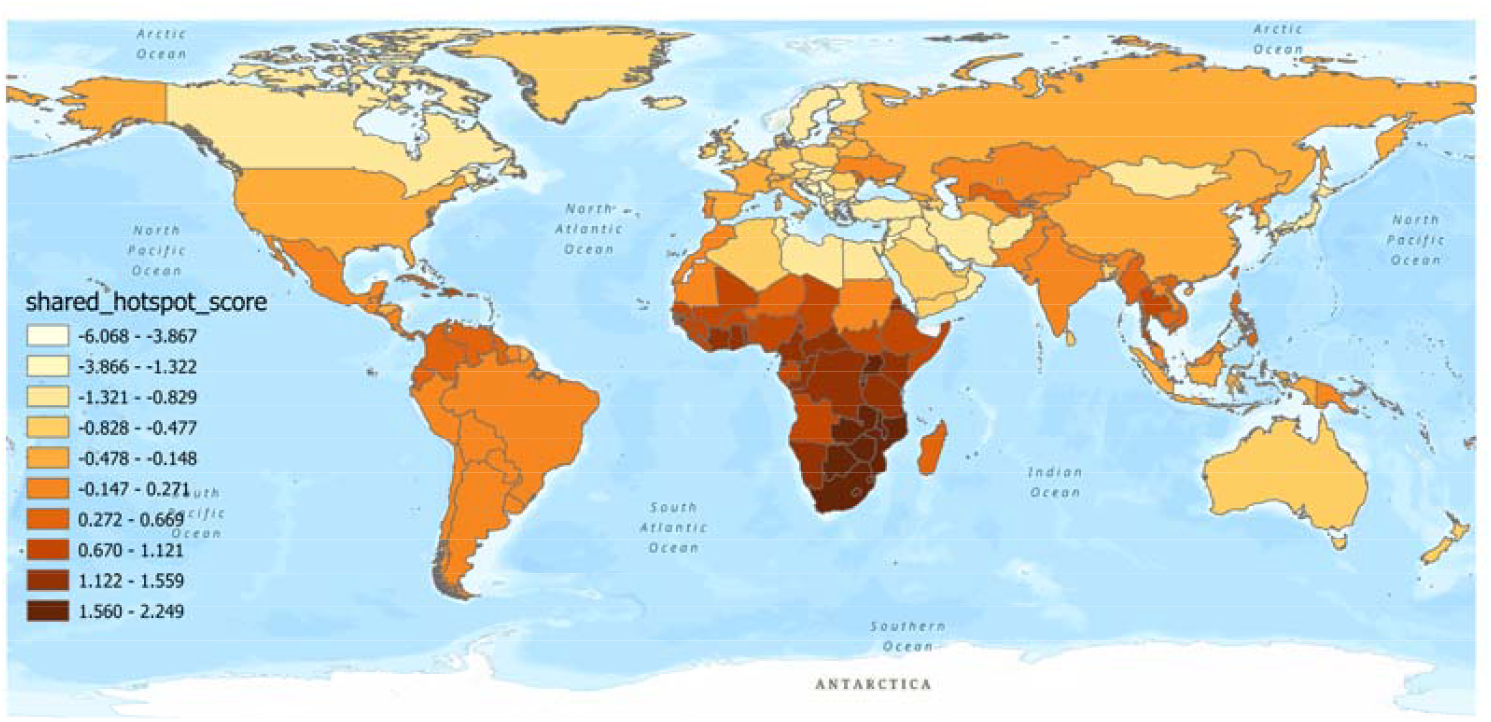
Global distribution of the shared spatial score (Note: The figures were created using ArcGIS.)

### 3.3. Posterior exceedance probability: hotspot stability

The posterior exceedance probability showed clear geographic differences in hotspot stability. The five countries with the strongest posterior support for being shared hotspots were **Botswana** (*P* = 1.000), **Central African Republic** (*P* = 1.000), **Burundi** (*P* = 1.000), **Zambia** (*P* = 1.000), and **Rwanda** (*P* = 1.000). More broadly, countries with high posterior support for hotspot status were concentrated mainly in **sub-Saharan Africa**, especially eastern and southern Africa, indicating that the shared hotspot pattern in this region was not only strong in magnitude but also highly stable.

In contrast, the countries least likely to be shared hotspots were **Albania** (*P* = 0.000), **Bosnia and Herzegovina** (*P* = 0.000), **Cook Islands** (*P* = 0.000), **Jordan** (*P*= 0.000), and **Montenegro** (*P*= 0.000). Low posterior exceedance probabilities were observed mainly in **southeastern Europe / the Balkans**, with additional very low-probability countries in some **Pacific island settings** and parts of the **Middle East**, indicating that these countries were highly unlikely to belong to the shared hotspot pattern(see Fig 5).

**Fig 5.**
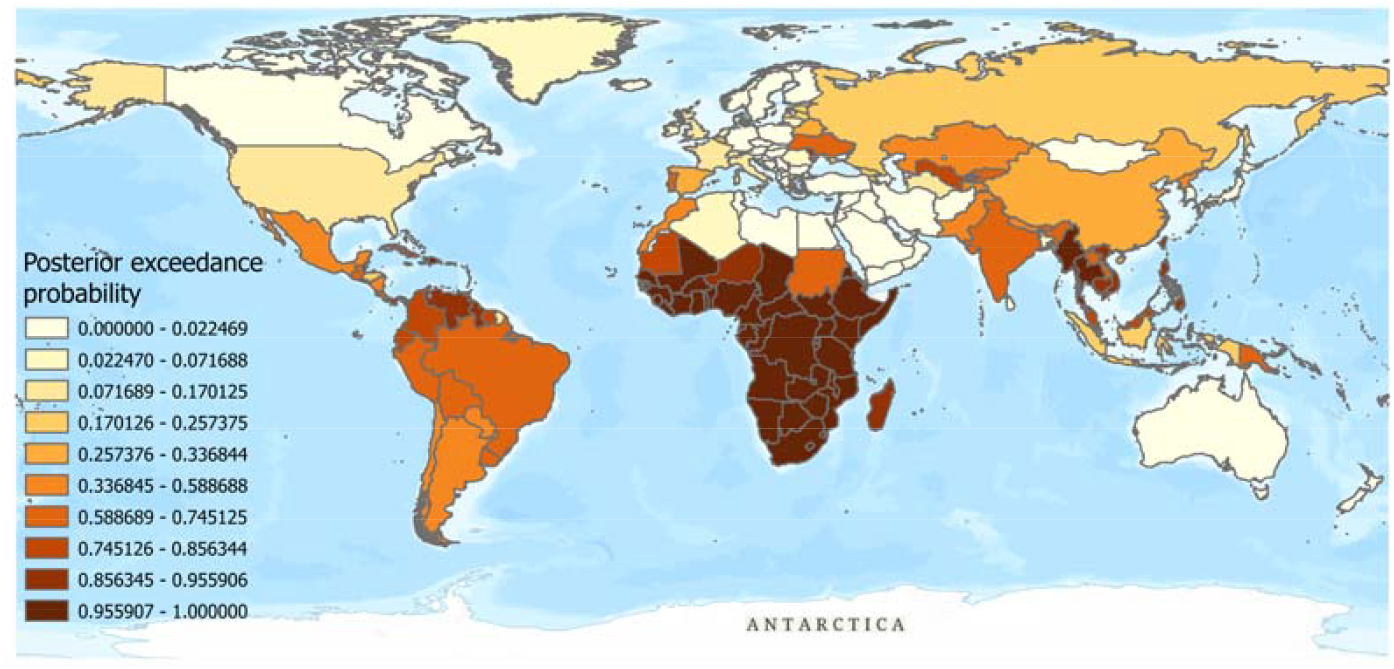
Global distribution of the posterior exceedance probability (Note: The figures were created using ArcGIS.)

### 3.4. Disease-specific patterns

Disease-specific loading patterns: The estimated disease-specific loadings showed clear differences in the extent to which each STI was aligned with the shared spatial hotspot pattern. **HIV/AIDS had the strongest loading on the shared component ()**, indicating that its spatial distribution was most strongly aligned with the common hotspot structure captured by the model. In contrast, **chlamydial infection had the weakest loading ()**, suggesting a comparatively smaller contribution from the shared spatial pattern. Syphilis showed an intermediate loading, indicating that it was influenced by the shared component to a greater extent than chlamydial infection but less strongly than HIV/AIDS. Overall, these results suggest that the shared hotspot structure was most strongly expressed in HIV/AIDS, while the degree of alignment with the shared pattern varied across the three STIs (see **Fig 6**).

**Fig 6.**
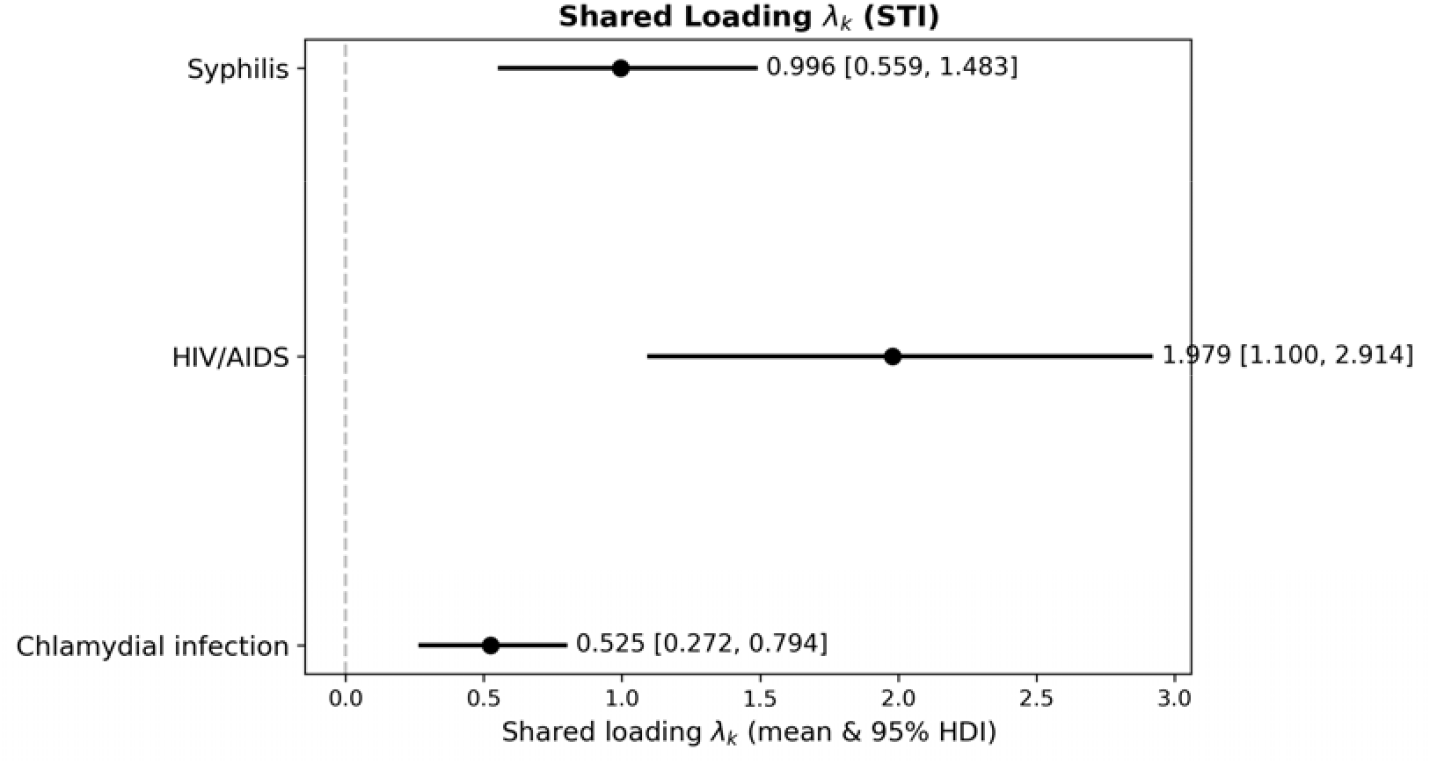
Disease-specific loadings on the shared spatial component (Note: The figures were created using Python.)

## 4. External validation

Detailed diagnostic procedures and full results are provided in **Supplementary Text S2**.

External validation supported the epidemiological plausibility of the shared spatial score. First, the score was strongly negatively correlated with SDI (*ρ*= −0.619, *p*= 6.38 × 10^-23^), indicating that higher shared spatial scores were concentrated in less socioeconomically developed settings. Second, the GEE analysis showed that a one-standard-deviation increase in the shared spatial score was associated with substantially higher HIV/AIDS mortality (IRR = 14.64, 95% CI: 11.90–18.01), while the upper-quartile group also had markedly higher mortality than the lower-quartile group (IRR = 315.39, 95% CI: 109.54–908.07). Together, these findings suggest that the shared spatial score captures meaningful epidemiological vulnerability.

## 5. Discussion

This study identified a clear and geographically structured shared spatial pattern across HIV/AIDS, syphilis, and chlamydial infection at the global level. Countries with the highest shared spatial scores were concentrated predominantly in sub-Saharan Africa, especially in eastern and southern Africa, whereas the lowest-scoring countries were more often observed in parts of southeastern Europe and a subset of Pacific island settings. The posterior exceedance probabilities further showed that the most stable positive shared spatial signals were also concentrated mainly in sub-Saharan Africa, indicating that this region was not only characterized by high shared spatial scores, but also by strong posterior support for persistent elevation in the common STI-related spatial pattern. In contrast, several countries in southeastern Europe, the Balkans, and selected Pacific settings showed consistently very low posterior exceedance probabilities, indicating that they were highly unlikely to belong to the positive side of the shared spatial pattern. Finally, the external validation analyses supported the epidemiological relevance of the shared spatial score, which was negatively associated with SDI and positively associated with HIV/AIDS mortality.

This is broadly consistent with previous studies showing that HIV/AIDS remains most heavily concentrated in sub-Saharan Africa and that syphilis also continues to impose a disproportionate burden in this region[10].

Beyond sub-Saharan Africa, several countries in mainland Southeast Asia and the Caribbean/northern South America also showed persistently positive shared spatial signals. In mainland Southeast Asia, Thailand (shared spatial score = 0.959, posterior exceedance probability = 0.998) and Myanmar (0.669, 0.978) were particularly notable, as both countries showed shared spatial scores in the moderate-to-upper range rather than among the global extremes, yet had very strong posterior support for a positive shared spatial pattern. A similar pattern was observed in the Caribbean and northern South America, including the Bahamas (0.965, 0.998), Barbados (0.844, 0.994), Tobago (0.838, 0.994), Jamaica (0.805, 0.993), Guyana (0.499, 0.925), Belize (0.485, 0.912), and Panama (0.483, 0.915). Haiti (1.423, 1.000) and the Dominican Republic (1.113, 1.000) showed stronger positive signals and were closer to the high-score end of the distribution. These findings indicate that, outside sub-Saharan Africa, some countries may not rank among the very highest in shared spatial score, but still exhibit highly stable posterior support for a persistently positive shared spatial structure.

Regarding the relationship between SDI and STI burden, our findings were broadly consistent with previous literature in suggesting that less socioeconomically developed or lower-resource settings tend to face greater burdens and control challenges related to HIV and other STIs[11].

From a public health perspective, the identified shared spatial pattern has important practical implications. Countries with persistently elevated shared spatial scores may reflect not merely the burden of a single STI, but broader weaknesses in the underlying prevention and control system. The co-occurrence of multiple STIs in the same settings suggests that common components of STI control—such as timely screening, effective treatment, partner notification, surveillance, and continuity of care—may be failing simultaneously. In this sense, the shared spatial score may serve as a quantitative signal of systemic vulnerability in STI prevention networks. It therefore provides a useful empirical basis for geographically prioritizing integrated STI services, particularly in settings where limited resources require more targeted deployment of screening, treatment, and public health outreach.

Methodologically, this study extends the application of shared-component spatial modelling to the field of sexually transmitted infections at the global level.

## 6. Limitations

Limitations. Several limitations should be acknowledged. First, the estimates of HIV/AIDS, syphilis, and chlamydial infection were derived from GBD data rather than directly observed surveillance counts, and therefore inherit the uncertainty of the underlying modelling process. Although GBD provides the most comprehensive and comparable country-level estimates currently available, measurement error and uncertainty in countries with limited primary data may still affect the precision of the inferred spatial pattern. Second, this was an ecological analysis conducted at the country level. As such, the identified associations should not be interpreted at the individual level, and no inference can be made about individual risk factors or individual-level co-occurrence of STIs. Third, some covariates contained substantial missingness and required imputation, particularly physician density, for which approximately 44% of values were missing. Although this approach allowed retention of global coverage, the imputed values may have introduced additional uncertainty into the adjusted estimates. Finally, the spatial resolution of the analysis was limited to the national level. This design cannot capture subnational heterogeneity, and important within-country variation in STI burden and service access may therefore have been obscured, especially in geographically large or highly unequal countries.

## 7. Conclusion

This study was undertaken in the context of WHO’s emphasis on people-centred and integrated STI prevention and care. By identifying the shared global spatial pattern of HIV/AIDS, syphilis, and chlamydial infection, our findings provide an evidence-based framework for the geographic prioritization of global STI prevention and control resources.

## Data Availability

The data used in this study were obtained from publicly available, aggregated, and de-identified sources. The original data sources are described in the manuscript. To prevent unintended disclosure of the processed files before final publication, the processed data, code, and supporting materials are currently stored in a private repository. Upon acceptance or formal publication of the manuscript, these materials will be deposited in a permanent public repository and made openly available without access restrictions.

https://docs.google.com/document/d/11xdRvnXTrmE3QREut7KKU6ptrs3odQ0x/edit

## 8. Data availability

To prevent unintended disclosure of the data while ensuring that the materials can be reviewed during peer review, the data and supporting files are currently stored in <u>a private repository</u>. Upon acceptance of the manuscript, the authors will deposit the data in a permanent public repository and make them openly available without access restrictions.

