## Supplementary Text 1 for "Bayesian shared-component spatiotemporal modeling of sexually transmitted infection co-occurrence: identifying geographic vulnerability across 204 countries, 1990–2023"

**Supplementary Text S1: Full Model Specification**

### 1.Likelihood

Let $y_{ikt}$denote the observed incidence count for disease $k$($k=1,\ldots,K$; $K=3$) in country $i$($i=1,\ldots,N$; $N=204$) and year $t$($t=1,\ldots,T$; $T=34$, corresponding to 1990–2023). We assumed[1]:

$$y_{ikt}\sim\text{NegativeBinomial}(\mu_{ikt},\text{ }\phi_{k}),$$

where $\mu_{ikt}$is the expected count and $\phi_{k}>0$is the disease-specific overdispersion parameter. Under this parameterisation, the variance is $\text{Var}(y_{ikt})=\mu_{ikt}+\mu_{ikt}^{2}/\phi_{k}$. As $\phi_{k}\to\infty$, the distribution converges to a Poisson. The negative binomial was chosen over the Poisson to accommodate the substantial overdispersion present in country-level STI incidence counts across 204 countries spanning multiple orders of magnitude.

**Prior:**

$$\phi_{k}\sim\text{Gamma}(2.0,\text{ }0.5),k=1,\ldots,K.$$

This prior has mean 4.0 and is moderately informative, placing most mass on values consistent with moderate-to-strong overdispersion while remaining compatible with near-Poisson behaviour.

### 2.Linear predictor

The logarithm of the expected count was modelled as[2]:

$$\log(\mu_{ikt})=\log(\text{Pop}_{it})+\alpha_{k}^{\left( 0 \right)}+\mathbf{x}_{it}^{\top}\boldsymbol{\gamma}_{k}+\lambda_{k}\text{ }s_{i}+\delta_{ki}+\theta_{kt},$$

where each component is described below.

#### 2.1.Population offset

$\log(\text{Pop}_{it})$is the natural logarithm of the total population of country $i$in year $t$, included as an offset so that the model effectively operates on incidence rates rather than raw counts.

#### 2.2.Disease-specific intercept

$\alpha_{k}^{\left( 0 \right)}$is the intercept for disease $k$, capturing the baseline (log) rate of disease $k$after accounting for all other model components.

**Prior:**

$$\alpha_{k}^{\left( 0 \right)}\sim\text{Normal}(0,\text{ }2.0),k=1,\ldots,K.$$

The relatively wide prior (standard deviation = 2.0 on the log-rate scale) accommodates the large differences in baseline incidence across the three STIs (e.g., chlamydia incidence is several orders of magnitude higher than HIV/AIDS incidence in many countries).

#### 2.3.Covariate fixed effects

$\mathbf{x}_{it}$is a $P$-dimensional vector ($P=5$) of time-varying covariates for country $i$in year $t$:

1. **SDI** (Socio-demographic Index): used on its original 0–1 scale;
2. **Log population density**: $\log(1+\text{pop\_density}_{it})$;
3. **Log health expenditure per capita**: $\log(1+\text{health\_pc}_{it})$;
4. **UHC service coverage index**: used on its original scale;
5. **Physician density** (per 1,000 population): used on its original scale.

All five covariates were z-score standardised (mean-centred and divided by standard deviation) across all country-year observations prior to model fitting, so that regression coefficients $\gamma_{kp}$are comparable in magnitude across covariates.

$\boldsymbol{\gamma}_{k}=(\gamma_{k1},\ldots,\gamma_{kP})^{\top}$is the vector of disease-specific regression coefficients.

**Prior:**

$$\gamma_{kp}\sim\text{Normal}(0,\text{ }1.0),k=1,\ldots,K;\text{ }p=1,\ldots,P.$$

A unit-scale prior on standardised coefficients implies that a one-standard-deviation change in a covariate is *a priori* expected to shift the log-rate by approximately ±1, which is weakly informative and allows the data to dominate.

#### 2.4.Shared spatial component

$s_{i}$is the shared spatial random effect for country $i$, representing the spatial pattern common to all three STIs. It is modelled using a scaled intrinsic conditional autoregressive (ICAR) prior[3].

**Construction.** We first draw a vector of unconstrained values and centre them:

$$s_{i}^{\text{raw}}\sim\text{Normal}(0,\text{ }1),i=1,\ldots,N,$$

$$\tilde{s}_{i}=s_{i}^{\text{raw}}-\bar{s}^{\text{raw}},$$

where $\bar{s}^{\text{raw}}=N^{-1}\sum_{i=1}^{N} s_{i}^{\text{raw}}$. The ICAR structure is imposed via a soft constraint (Potential):

$$\log p(\tilde{\mathbf{s}})+=-\frac{1}{2\tau_{s}}\sum_{i\sim j} (\tilde{s}_{i}-\tilde{s}_{j})^{2},$$

where the sum runs over all pairs of neighbouring countries ($i\sim j$), and $\tau_{s}$is the ICAR scaling factor (see below).

##### 2.4.1.ICAR scaling factor

To ensure that the marginal variance of the spatial field is interpretable regardless of the specific neighbourhood structure, we computed a scaling factor $\tau_{s}$following the approach of Sørbye and Rue (2014)[4]. Specifically, let $\mathbf{Q}$denote the $N\times N$structure matrix of the adjacency graph, with $Q_{ii}=n_{i}$(the number of neighbours of country $i$), $Q_{ij}=-1$if $i\sim j$, and $Q_{ij}=0$otherwise. Because the ICAR model is rank-deficient (the constant vector is in the null space), we computed the Moore–Penrose generalised inverse $\mathbf{Q}^{-}$via eigendecomposition, retaining only positive eigenvalues ($>{10}^{-10}$). The scaling factor was then defined as the geometric mean of the marginal variances:

$$\tau_{s}=\exp\text{ }\text{,}\left( \frac{1}{N}\sum_{i=1}^{N} \log[\mathbf{Q}^{-}]_{ii} \right).$$

**Scaled shared spatial effect.** The final shared spatial component entering the linear predictor is:

$$s_{i}=\frac{\sigma_{s}}{\sqrt{\tau_{s}}}\text{ }\tilde{s}_{i},$$

where $\sigma_{s}>0$controls the overall magnitude of the shared spatial variation.

**Priors:**

$$\sigma_{s}\sim\text{HalfNormal}(1.0).$$

#### 2.5.Disease-specific loading

$\lambda_{k}\geq0$is the non-negative loading coefficient that links disease $k$to the shared spatial pattern. Its magnitude reflects how strongly disease $k$tracks the common spatial structure; a larger $\lambda_{k}$implies that disease $k$is more tightly coupled to the shared pattern[2].

**Prior:**

$$\lambda_{k}\sim\text{HalfNormal}(1.0),k=1,\ldots,K.$$

The HalfNormal prior constrains $\lambda_{k}\geq0$to ensure identifiability of the shared component's sign.

#### 2.6.Disease-specific spatial deviation

$\delta_{ki}$captures spatial variation specific to disease $k$in country $i$, beyond what is explained by the shared spatial component. This allows each disease to exhibit its own residual geographic pattern.

**Prior:**

$$\delta_{ki}\sim\text{Normal}(0,\text{ }\sigma_{\delta}),k=1,\ldots,K;\text{ }i=1,\ldots,N,$$

$$\sigma_{\delta}\sim\text{HalfNormal}(1.0).$$

Note that $\sigma_{\delta}$is shared across all diseases and countries (a single global scale parameter), providing regularisation and partial pooling across the disease-specific spatial deviations.

#### 2.7.Disease-specific temporal effect

$\theta_{kt}$is the temporal random effect for disease $k$at time $t$, modelled as a first-order random walk [RW(1)] with non-centred parameterisation and a sum-to-zero constraint[5].

**Construction.** For each disease $k$, we draw $T-1$innovation terms:

$$\epsilon_{k,t}\sim\text{Normal}(0,\text{ }1),t=1,\ldots,T-1,$$

and construct the cumulative sum:

$$\theta_{k,t}^{\text{raw}}=\left\{ \begin{matrix} 0 & \text{if }t=1, \\ \sum_{j=1}^{t-1} \sigma_{\text{rw},k}\cdot\epsilon_{k,j} & \text{if }t=2,\ldots,T. \end{matrix} \right.$$

A sum-to-zero constraint is then applied for identifiability (to avoid confounding with the intercept):

$$\theta_{kt}=\theta_{k,t}^{\text{raw}}-\frac{1}{T}\sum_{t^{'}=1}^{T} \theta_{k,t^{'}}^{\text{raw}}.$$

This non-centred parameterisation (sampling standardised innovations $\epsilon_{k,t}$rather than the $\theta_{kt}$directly) improves MCMC sampling efficiency for random walk models, particularly when the innovation variance $\sigma_{\text{rw},k}^{2}$is small, as it reduces the funnel-shaped geometry that can cause divergent transitions in Hamiltonian Monte Carlo.

Prior on innovation scale:

$$\sigma_{\text{rw},k}\sim\text{HalfNormal}(0.5),k=1,\ldots,K.$$

The prior with scale 0.5 reflects the expectation that year-to-year changes in the log-rate are modest (i.e., smooth temporal trends).

### 3.Spatial adjacency structure

Country-level spatial adjacency was defined using Queen contiguity from a Natural Earth 10 m resolution shapefile, where two countries are neighbours if they share any boundary point. For island nations and geographically isolated territories with no contiguous neighbours ($n=52$ countries), we applied a K-nearest neighbours (KNN) fallback with $K=5$, based on great-circle distances between country centroids. KNN edges were made symmetric: if country $i$was added as a neighbour of country $j$, then $j$was also added as a neighbour of $i$.

### 4.Covariate missingness handling

Missing covariate values were imputed using a three-step sequential procedure:

1. **Within-country temporal interpolation:** for each country, missing values were filled by forward-fill followed by backward-fill along the time axis;
2. **Cross-country year-level imputation:** remaining missing values were replaced by the median value across all countries within the same year;
3. **Global fallback:** any remaining missing values were filled with the global median across all observations.

The proportion of missing values before imputation was: population (2.0%), SDI (0.0%), population density (4.1%), health expenditure per capita (34.8%), UHC index (33.2%), and physician density (44.3%). A missingness report is provided in Supplementary Table S2. The relatively high missingness rates for health expenditure, UHC, and physician density are acknowledged as a limitation; a complete-case sensitivity analysis could be considered in future work.

### 5.MCMC sampling configuration

Parameter estimation was performed using the No-U-Turn Sampler (NUTS), a variant of Hamiltonian Monte Carlo, as implemented in PyMC. The sampling configuration was showed in Table 1 [6, 7]:

Table 1 MCMC sampling configuration

| Parameter | Value |
| --- | --- |
| Number of chains | 16 |
| Tuning (warm-up) iterations per chain | 2,000 |
| Post-warm-up draws per chain | 2,000 |
| Total posterior samples | 32,000 |
| Target acceptance probability | 0.97 |
| Maximum tree depth | 15 |
| Random seed | 42 |

The elevated target acceptance probability (0.97, compared to the default of 0.80) and maximum tree depth (15, compared to the default of 10) were necessary to navigate the complex posterior geometry arising from the ICAR spatial structure over 204 countries. Log-likelihood values were computed during sampling to enable subsequent LOO-CV and WAIC model comparison without re-fitting[8].

### 6.Summary of all priors

For clarity, all prior distributions used in the main model are summarised in Table 2

Table 2 Summary of prior distributions used in the main model

| **Parameter** | **Prior** | **Dimension** | **Description** |
| --- | --- | --- | --- |
| $\alpha_{k}^{\left( 0 \right)}$ | $\text{Normal}(0,2.0)$ | $K$ | Disease-specific intercept |
| $\gamma_{kp}$ | $\text{Normal}(0,1.0)$ | $K\times P$ | Covariate coefficients |
| $\sigma_{\text{rw},k}$ | $\text{HalfNormal}(0.5)$ | $K$ | RW(1) innovation scale |
| $\epsilon_{k,t}$ | $\text{Normal}(0,1)$ | $K\times(T-1)$ | RW(1) innovations (non-centred) |
| $s_{i}^{\text{raw}}$ | $\text{Normal}(0,1)$ | $N$ | Shared spatial (unconstrained) |
| $\sigma_{s}$ | $\text{HalfNormal}(1.0)$ | 1 | Shared spatial scale |
| $\lambda_{k}$ | $\text{HalfNormal}(1.0)$ | $K$ | Disease-specific spatial loading |
| $\sigma_{\delta}$ | $\text{HalfNormal}(1.0)$ | 1 | Disease-specific spatial deviation scale |
| $\delta_{ki}$ | $\text{Normal}(0,\sigma_{\delta})$ | $K\times N$ | Disease-specific spatial deviation |
| $\phi_{k}$ | $\text{Gamma}(2.0,0.5)$ | $K$ | Negative binomial overdispersion |

where $K=3$(diseases), $N=204$(countries), $T=34$(years), $P=5$(covariates).

### 7.Shared spatial score

The primary output of the model is the country-level shared spatial score, defined as the posterior mean of the shared spatial component:

$$\bar{s}_{i}=E(s_{i}\mid\mathbf{y}),i=1,\ldots,N,$$

estimated as the Monte Carlo average over $S=32,000$posterior samples. This is a continuous index summarising the country-specific latent spatial pattern shared across HIV/AIDS, syphilis, and chlamydial infection after adjusting for covariates, disease-specific spatial effects, and disease-specific temporal effects. Because the shared spatial component is centered, a positive score indicates a higher-than-reference shared spatial tendency, whereas a negative score indicates a lower-than-reference shared spatial tendency; the magnitude reflects the strength of deviation on the log-rate scale. This score was used for ranking countries and for downstream external validation analyses.

### 8.Posterior stability

To assess the reliability of each country’s score direction, we computed the posterior exceedance probability:

$$P(s_{i}>0\mid\mathbf{y})=\frac{1}{S}\sum_{m=1}^{S} \mathbf{1}(s_{i}^{\left( m \right)}>0),$$

where $s_{i}^{\left( m \right)}$denotes the $m$-th posterior sample of $s_{i}$. Values close to 1.0 or 0.0 indicate that the sign of the shared spatial score is stable across the posterior distribution, whereas values near 0.5 indicate substantial uncertainty about whether the country’s shared spatial tendency lies above or below the centered reference level. For descriptive purposes, countries with $P(s_{i}>0\mid\mathbf{y})>0.95$are referred to as “high-confidence hotspots” and those with $P(s_{i}>0\mid\mathbf{y})<0.05$as “high-confidence coldspots,” although emphasis throughout is placed on the continuous score and its posterior stability rather than on binary classification.
