## Supplementary Text S2 for "Bayesian shared-component spatiotemporal modeling of sexually transmitted infection co-occurrence: identifying geographic vulnerability across 204 countries, 1990–2023"

**Supplementary Text S2: Model Diagnostics and Validation Results**

### 1.MCMC Convergence

#### 1.1 Method:

Convergence of the No-U-Turn Sampler was assessed using three standard criteria: (i) the potential scale reduction factor R-hat, which compares between-chain and within-chain variance and should be close to 1.0 for well-mixed chains; (ii) the effective sample size (ESS), which quantifies the number of effectively independent posterior draws after accounting for autocorrelation; and (iii) the number of divergent transitions, which indicate regions of the posterior geometry that the sampler failed to explore adequately. We required R-hat ≤ 1.01 for all parameters, minimum bulk ESS > 400, and zero divergent transitions.

#### 1.2 Results**:**

All convergence criteria were satisfied (Table 1). The maximum R-hat across all model parameters was 1.0018, well below the 1.01 threshold, with zero parameters exceeding 1.01 or 1.05. The minimum bulk ESS was 9,853, far exceeding the 400 minimum, indicating that autocorrelation was negligible. Zero divergent transitions were recorded out of 32,000 total post-warmup samples (16 chains × 2,000 draws). The worst-performing parameter was spatial_disease[2, 48], which nonetheless had R-hat = 1.0018 and bulk ESS = 9,853—both indicating excellent mixing. These results confirm that the sampler thoroughly explored the posterior distribution.

Table 1 MCMC convergence summary

| Diagnostic | Value |
| --- | --- |
| Maximum R-hat | 1.0018 |
| Parameters with R-hat > 1.01 | 0 |
| Parameters with R-hat > 1.05 | 0 |
| Worst R-hat parameter | spatial_disease[2, 48] |
| Minimum bulk ESS | 9,853 |
| Parameters with bulk ESS < 400 | 0 |
| Minimum tail ESS | 15,348 |
| Divergent transitions | 0 / 32,000 (0.0%) |
| Total post-warmup samples | 32,000 |

Note：The parameter with the largest R-hat was one element of the disease-specific spatial effect matrix (spatial_disease [2,48]), but its R-hat was still only 1.0018.

### 2. Model Fit: LOO-CV and WAIC

#### 2.1Method

Model fit was assessed using two information criteria based on the log-pointwise-predictive-density: leave-one-out cross-validation (LOO-CV) estimated via Pareto-smoothed importance sampling and the widely applicable information criterion [1, 2]. Both are computed from the 32,000 posterior samples of the log-likelihood matrix (20,808 observations).

The Pareto k diagnostic was used to identify observations for which the importance sampling approximation may be unreliable: k ≤ 0.70 indicates a reliable estimate ("good"), 0.70 < k ≤ 1.0 indicates a potentially unreliable estimate ("bad"), and k > 1.0 indicates a very unreliable estimate[3].

#### 2.2Results

The LOO and WAIC estimates were highly consistent (Table 2). Critically, 100% of observations (20,808/20,808) had Pareto k ≤ 0.70 ("good"), indicating that no single observation exerted undue influence on the model fit and that the importance sampling approximation underlying LOO-CV was reliable for all observations. The effective number of parameters (p_LOO = 623; p_WAIC = 622) is plausible for a model with 204 countries × 3 diseases over 34 years plus shared and disease-specific spatial and temporal random effects.

Table 2 Model comparison via LOO-CV and WAIC

| Metric | ELPD | SE | p_eff |
| --- | --- | --- | --- |
| LOO-CV | −176,314 | 471 | 623 |
| WAIC | −176,313 | 471 | 622 |

Table 3 Pareto k diagnostics:

| Category | Count | Percentage |
| --- | --- | --- |
| (−∞, 0.70] (good) | 20,808 | 100.0% |
| (0.70, 1.0] (bad) | 0 | 0.0% |
| (1.0, ∞) (very bad) | 0 | 0.0% |

### 3.Posterior Predictive Checks

#### 3.1 Method

Posterior predictive checks (PPC) were conducted by drawing 300 replicated datasets from the posterior predictive distribution and comparing them against observed data. Three types of checks were performed: (i) marginal density comparison of log(1 + y) between observed and replicated counts; (ii) scatter plots of observed versus posterior predictive mean at the observation level; and (iii) country-level time series comparisons for a sample of 10 countries (5 highest-burden and 5 randomly selected), showing observed counts against the posterior predictive median and 90% prediction intervals, stratified by disease.

#### 3.2 Results

The marginal posterior predictive distribution closely matched the observed distribution of log-transformed counts for all three diseases (see Fig 1).

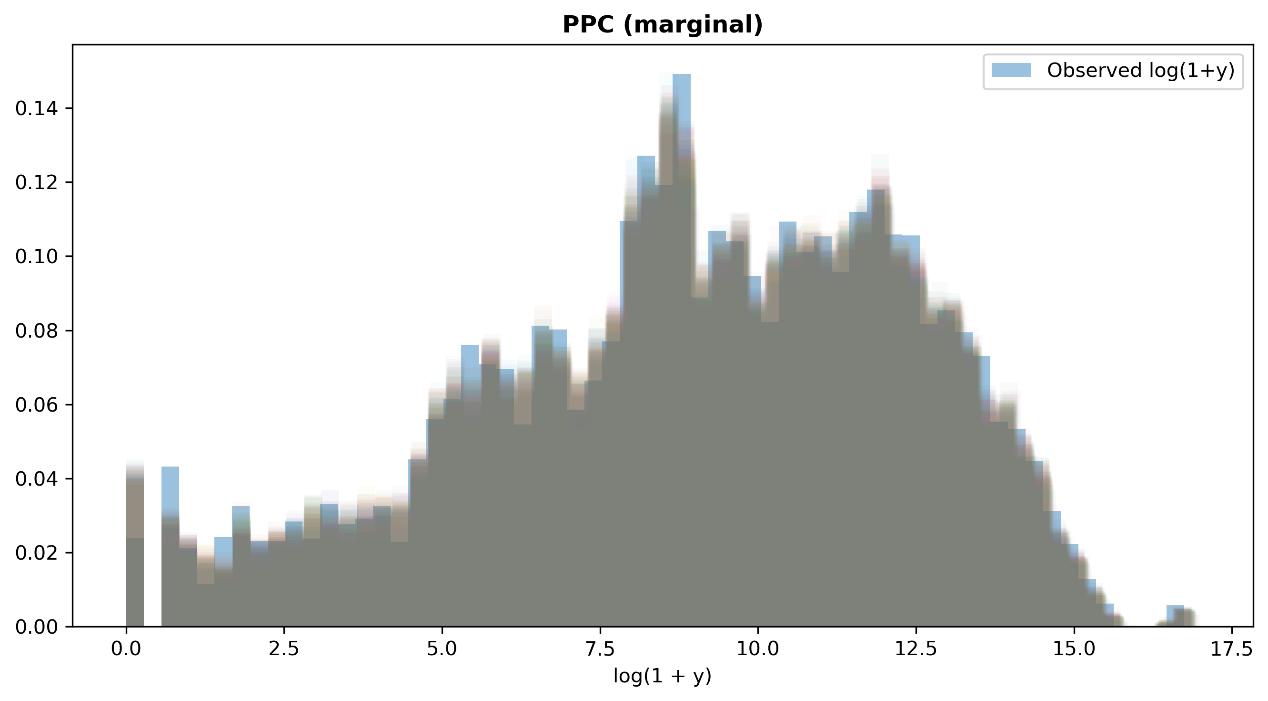

Fig 1 Marginal posterior predictive check for log-transformed counts

The marginal posterior predictive distribution closely overlapped the observed distribution of log-transformed counts, indicating that the fitted model reproduced the overall distributional shape of the data reasonably well. No major discrepancy was evident across most of the support of the distribution.

Scatter plots of observed versus posterior predictive means showed strong agreement along the diagonal with no systematic bias (see Fig 2).

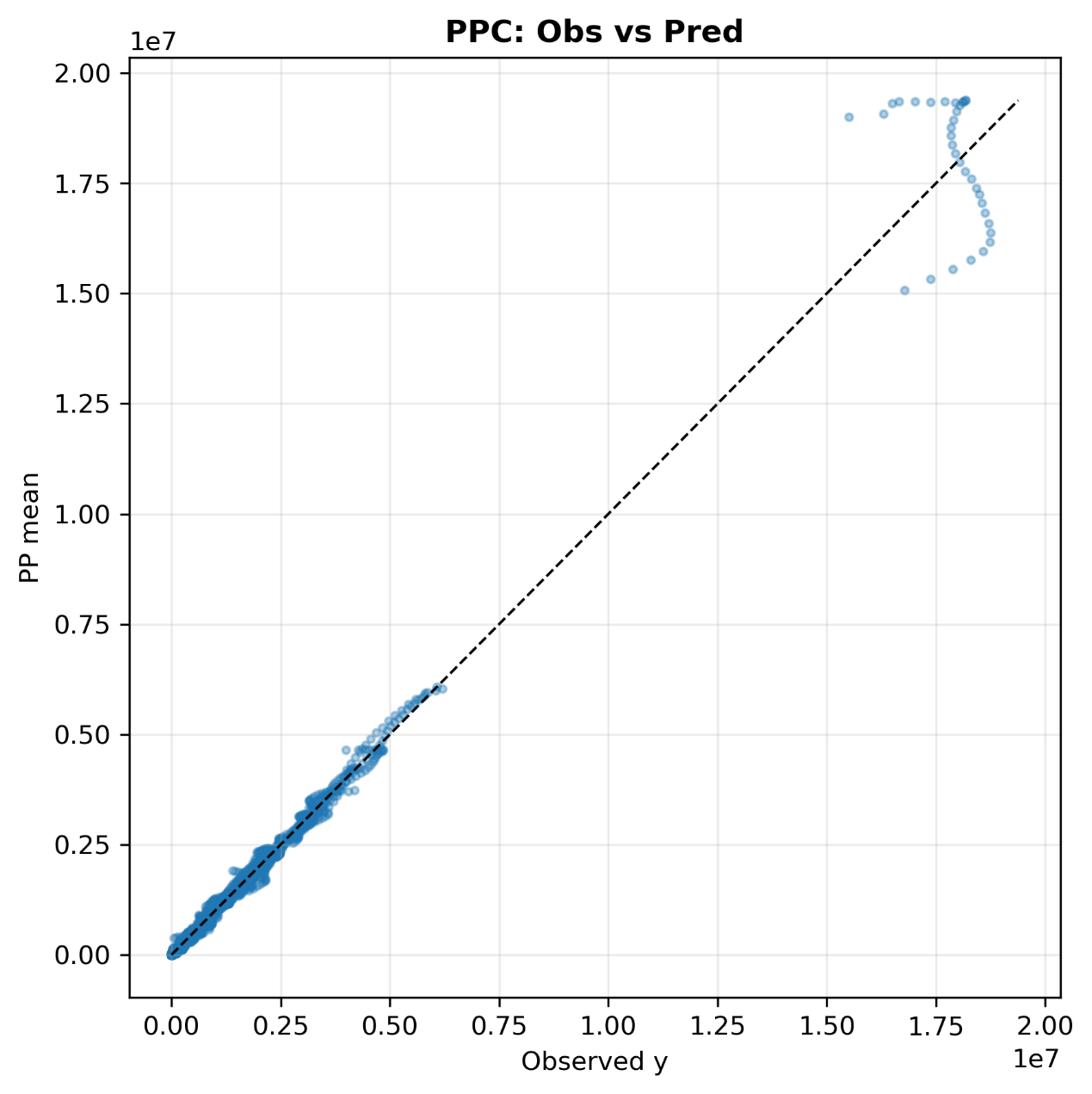

Fig 2 Observation-level posterior predictive check: observed values versus posterior predictive means

The scatter plot of observed values versus posterior predictive means showed close agreement along the 45-degree reference line for the majority of observations, indicating that the model reproduced the observation-level mean structure reasonably well. Some deviation was evident at the extreme upper end of the distribution, suggesting that the fit for the largest observed counts was less precise than for the bulk of the data.

Country-level posterior predictive checks showed that the observed time series generally fell within the 90% posterior predictive intervals across the selected countries and diseases, indicating that the model captured the main temporal patterns reasonably well. The posterior predictive medians broadly tracked the long-term trends of the observed counts, although some deviations were evident for abrupt peaks or turning points in specific country-disease series. Overall, these plots did not suggest systematic model misspecification at the country-year level. (See Fig 3-Fig 5).

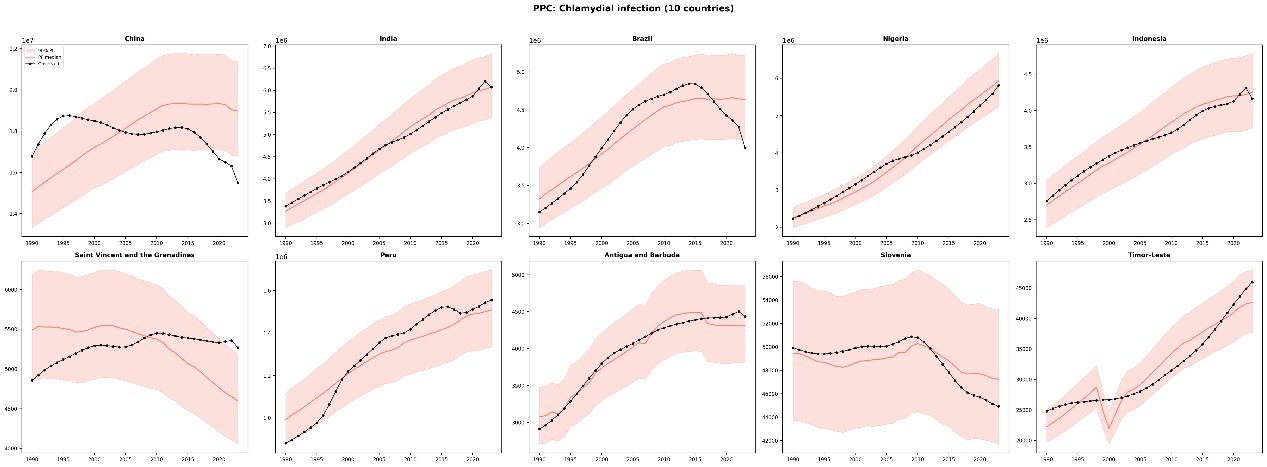

Fig 3 Country-level posterior predictive checks for Chlamydial infection

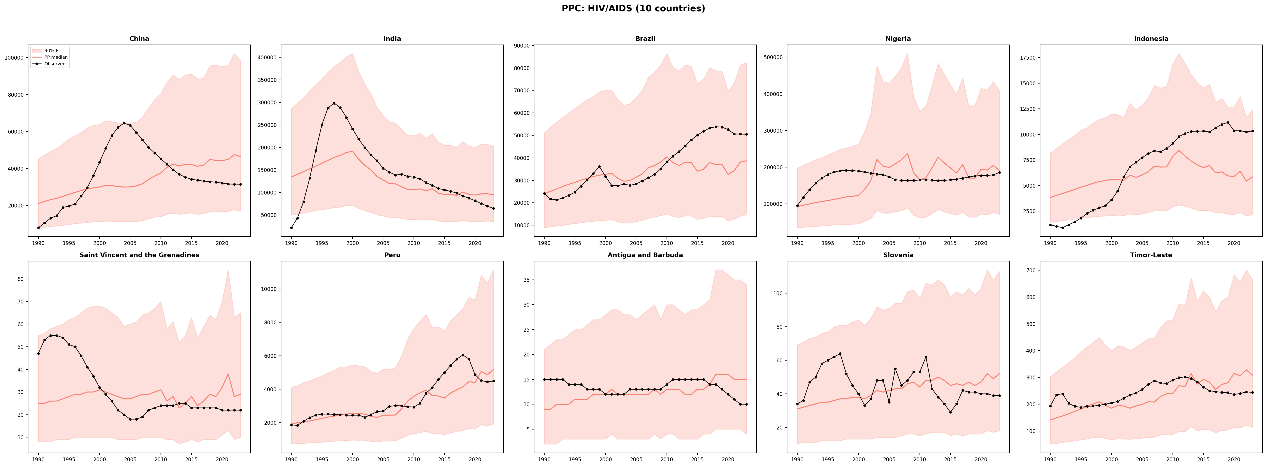

Fig 4 Country-level posterior predictive checks for HIV/AIDS

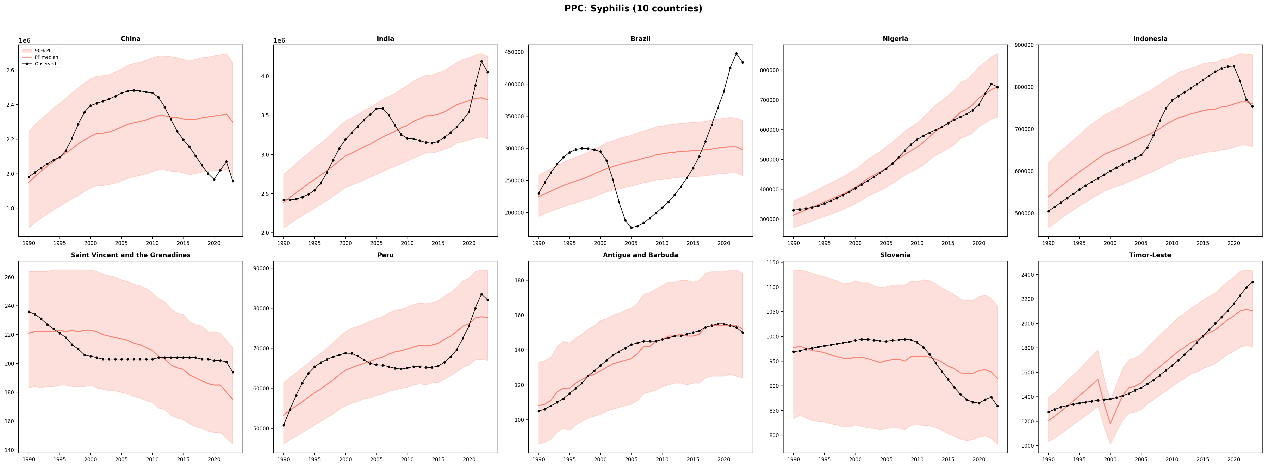

Fig 5 Country-level posterior predictive checks for Syphilis

### 4. Prior Sensitivity Analysis

#### 4.1 Method

To assess the robustness of the shared hotspot rankings to prior choices, the model was re-fitted under two alternative prior configurations, each modifying one prior while holding all others at their baseline values(see Table 4):

Table 4 Alternative prior configurations used in the prior sensitivity analysis

| Configuration | Modified prior | Baseline | Alternative |
| --- | --- | --- | --- |
| alpha_nb_alt | Overdispersion φ_k | Gamma(2.0, 0.5) | Gamma(1.0, 0.1) |
| sigma_s_alt | Shared spatial scale σ_s | HalfNormal(1.0) | HalfNormal(2.0) |

Stability was quantified using: (i) Spearman rank correlation between country-level shared spatial scores under each alternative versus the baseline configuration; and (ii) the overlap (out of 10) of the top-10 hotspot countries.

#### 4.2Results

The shared hotspot rankings were essentially invariant to both alternative prior specifications (see Table 5). Spearman correlations exceeded 0.9999 for both alternatives, and the top-10 hotspot countries were identical across all three configurations. These results indicate that the posterior is overwhelmingly dominated by the data rather than the prior, consistent with the large sample size (20,808 observations across 204 countries and 34 years).

Table 5 Prior sensitivity analysis results

| Configuration | Spearman ρ vs baseline | Top-10 overlap |
| --- | --- | --- |
| Baseline | 1.0000 | 10 / 10 |
| alpha_nb_alt | 0.9999 | 10 / 10 |
| sigma_s_alt | 0.9999 | 10 / 10 |

### 5. Covariate Collinearity

#### 5.1Method

Collinearity among the five covariates was assessed using: (i) Spearman rank correlation matrix; and (ii) variance inflation factors (VIF), where VIF > 10 is conventionally considered indicative of problematic multicollinearity.

#### 5.2Results

No covariate exhibited a VIF exceeding 10 (see Table 6). The highest VIF was 6.31 for SDI, reflecting its known correlation with health expenditure (Spearman ρ = 0.72) and UHC coverage (ρ = 0.68). While this value is moderately elevated, it remains below the conventional threshold of 10 and is typical for socioeconomic indicators in global ecological analyses. All other covariates had VIF below 5.

Table 6 Variance inflation factors for model covariates

| Covariate | VIF |
| --- | --- |
| SDI | 6.31 |
| Log population density | 1.04 |
| Log health expenditure per capita | 4.50 |
| UHC service coverage index | 4.59 |
| Physician density per 1,000 | 2.22 |

The Spearman correlation matrix showed strong positive correlations among SDI, health expenditure, UHC coverage, and physician density, whereas log population density was only weakly correlated with the other covariates. This pattern indicates that the socioeconomic and health-system indicators capture related dimensions, but the subsequent VIF results suggested that the degree of multicollinearity remained within an acceptable range (see Fig 6).

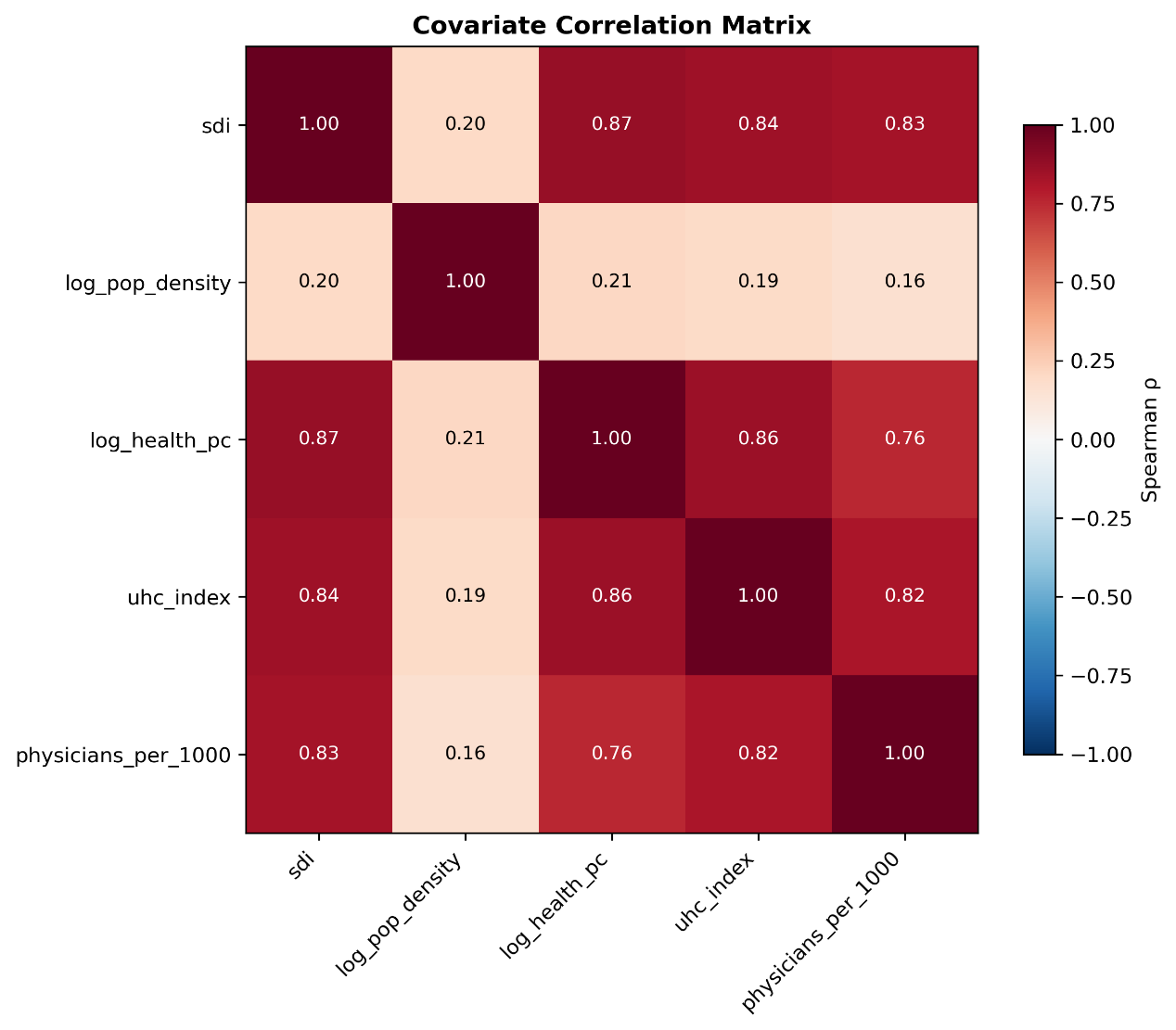

Fig 6. Spearman correlation matrix of model covariates

### 6.External Validation 1: Convergent Validity with SDI

#### 6.1Method

To assess whether the identified shared hotspots are consistent with known patterns of socioeconomic development, we computed the Spearman rank correlation between country-level shared spatial scores and the most recent available SDI values (2023). The expected direction is negative: countries with higher SDI (indicating greater socioeconomic development) should have lower STI co-occurrence burden, reflecting stronger health systems, greater access to screening and treatment, and more effective prevention programmes.

#### 6.2Results

A strong and highly significant negative correlation was observed (see Table 7), consistent with the hypothesised direction. Countries identified as shared hotspots overwhelmingly had low SDI values, while shared coldspots had high SDI values.

Table 7 SDI convergent validity

| Statistic | Value |
| --- | --- |
| N (countries) | 204 |
| Spearman ρ | −0.619 |
| p-value | 6.38 × 10⁻²³ |
| SDI year | 2023 |
| Expected direction | Negative (confirmed) |

### 7. Convergent Validity with HIV/AIDS Mortality

#### 7.1 **Method**

The biological and epidemiological rationale for this validation is that HIV/AIDS shares sexual transmission routes with the three modelled STIs, and STI co-occurrence—particularly genital ulcerative diseases such as syphilis—is known to facilitate HIV transmission. Thus, countries with high shared STI spatial burden should exhibit elevated HIV/AIDS mortality. We fitted a GEE negative binomial model with exchangeable within-country working correlation, regressing HIV/AIDS death counts (GBD 2023, 204 countries, 1990–2023, 6,936 observations) on the shared spatial score with log(population) as offset, year fixed effects, and the same five z-score standardised covariates used in the main model. Country fixed effects were not included because the shared spatial score is time-invariant and would be absorbed. Two models were fitted: (i) a main model with the continuous shared spatial score; and (ii) a sensitivity model comparing the upper quartile (high co-occurrence) versus lower quartile (low co-occurrence) of the hotspot score distribution.

#### 7.2 Results

Both models confirmed a strong positive association between STI co-occurrence and HIV/AIDS mortality (see Table 8).

Table 8 GEE convergent validity: HIV/AIDS mortality

| Model | Exposure | IRR | 95% CI | p-value | N (obs) | N (countries) |
| --- | --- | --- | --- | --- | --- | --- |
| Main (continuous) | Shared spatial score (z) | 14.64 | 11.90–18.01 | 6.35 × 10⁻¹⁴² | 6,936 | 204 |
| Sensitivity (categorical) | High vs Low group | 315.39 | 109.54–908.07 | 1.50 × 10⁻²⁶ | 3,468 | 102 |

In the main model, a one-standard-deviation increase in the shared spatial score was associated with a 14.6-fold increase in HIV/AIDS mortality rate (IRR = 14.64, 95% CI: 11.90–18.01). Among the covariates, UHC coverage was protective (IRR = 0.60, p < 0.001), consistent with the expectation that higher health service coverage reduces HIV/AIDS mortality.

In the sensitivity model, the upper-quartile co-occurrence group had HIV/AIDS mortality rates over 300 times higher than the lower-quartile group after adjustment for covariates and year effects. While this effect size is very large, it reflects the enormous heterogeneity in HIV/AIDS mortality across countries—sub-Saharan African hotspot countries have HIV/AIDS death rates several orders of magnitude higher than low-burden coldspot countries in Western Europe and East Asia.
